# Functional Capacity and Physical Activity in COPD: Associations with Hospitalizations, Health related Quality of Life and Mortality. A Prospective study

**DOI:** 10.64898/2026.01.04.26343342

**Authors:** Cristóbal Esteban, Irantzu Barrio, Amaia Aramburu, Abelardo Monsalve-Cobis, Leyre Chasco, Javier Moraza, Josu Najera-Zuloaga, Susana Aizpiri, Myriam Aburto, Inmaculada Arostegui, José M. Quintana

## Abstract

**Objective:** To demonstrate the association between hospital admissions, health-related quality of life (HRQoL), limitations in daily activities, mortality, and survival in relation to physical activity (PA) and functional capacity (FC), which were used together to classify a cohort of COPD patients.

**Methods:** Prospective study. Participants were consecutively recruited from six outpatient respiratory clinics of a single university hospital. Data were recorded on sociodemographic and clinical variables, dyspnea (Medical Research Council), HRQoL (Saint George’s Respiratory Questionnaire (SGRQ) and COPD Assessment Test (CAT)), and limitation of daily life (London Chest Activity of Daily Living scale (LCADL)). FC was evaluated with a 6-minute walk test. PA (average daily steps) was measured using an accelerometer. Participants were grouped into four “CanDo/DoDo” quadrants based on their PA and FC combinations.

**Results:** 401 patients were included. Compared to the “Can/Do” group, the “Can’t/Do” (Odds Ratio (OR): 3.34) and the “Can’t/Don’t” (OR: 27.3) groups had a higher risk of dying within one year, worse survival rates (Hazard Ratios: 2.37 and 2.87), and a higher risk of hospital admission (OR: 2.83 and 2.84) and additional admissions (Incidence Rate Ratios of 2.58 and 2.31) in the first year of follow-up. The “Can’t/Do” and the “Can’t/Don’t” groups had worse scores on the SGRQ (OR: 1.83 and 2.15), CAT (OR: 2.44 and 3.39), and LCADL (OR: 1.31 and 1.42) questionnaires conducted at the beginning of the study.

**Conclusion:** Using PA and FC quadrants to classify participants showed associations with robust outcomes, thus enabling strategies to be developed to improve those outcomes.

## INTRODUCTION

There is no doubt that physical activity (PA) is beneficial for health; however, from an evolutionary perspective, PA is currently viewed in a very different context compared to that of our hunter-gatherer ancestors [1]. PA and functional capacity (FC)—defined as the functioning of several integrated systems to fulfill a task—are two key elements for health; therefore, attaining the highest level in both has significant consequences throughout life.

A new medical approach needs to be designed that focuses not just on lifespan but also on “healthspan,” with the aim of reducing the gap between the two, not only among the general population but particularly among individuals with chronic diseases [2]. In fact, multimorbidity is the usual situation in chronic diseases, particularly associated with aging [3], which leads to impairments in the overall condition of these patients. However, what patients value most is maintaining their independence [4]. In this scenario, PA and FC could play an essential role, assuming that maintaining independence requires a multidimensional approach [5].

Generally speaking, having adequate FC means adhering to an established PA program, allowing several systems to work together to achieve a single objective. In contrast, engaging in PA primarily involves behavioral attitudes. In other words, while FC and PA are related, they encompass different constructs [6]. However, these general distinctions can prove simplistic when other areas are taken into consideration. In this regard, Bauman et al. studied the relationship between environmental factors and PA through their “ecological model” [7]. Applying this model to COPD has revealed a negative association between PA and factors such as population density, terrain gradient, and annual average residential concentrations of NO2. Interestingly, an association has also been found between certain environmental factors and FC, demonstrating both positive and negative relationships with terrain gradient and population density, respectively [8]. Additionally, there have been noteworthy findings in PA concerning its relationship with genetic factors [9,10]. Therefore, everything appears to be interconnected, and many intriguing questions require exploration regarding the underlying mechanisms involved.

In a significant proposal, Vaes et al. recently recommended classifying people with COPD based on their FC and PA [11]. The association between low levels of PA and several chronic diseases is well established [12]. Maintaining FC has been identified as one of the key elements for mitigating the limiting effects of disease progression and ensuring healthy aging, thereby extending healthspan through secondary prevention [13].

The aim of this study was to demonstrate the association between hospital admissions, health-related quality of life (HRQoL), limitations in daily activities, mortality, and survival in relation to PA and FC, which were used together to classify a cohort of individuals with COPD, with a view to proposing potential therapeutic strategies for improving PA and FC to ameliorate these outcomes.

## METHODS

### Participants and data collection

Prospective, cohort study. Participants were recruited at one of six outpatient respiratory clinics run by the Respiratory Service of Galdakao University Hospital. The functional criteria for inclusion were forced expiratory volume in one second (FEV1) <80% of the predicted value and a FEV1/forced vital capacity ratio <70%. Participants were consecutively included in the study if they had been diagnosed with COPD for at least 6 months and had been stable for at least 6 weeks. Participants were not deemed eligible for the study if they had been diagnosed with asthma, any other major respiratory diseases, or psychiatric or neurological problems that might hinder effective collaboration. The protocol was approved by the hospital’s ethics and research committees (16/2014). All candidate patients were given detailed information about the study and all those included provided written informed consent.

### Study protocol

Sociodemographic, and clinical variables were recorded. Dyspnea was categorized using the modified Medical Research Council (mMRC) dyspnea scale [14]. Comorbidities were summarized using the Charlson comorbidity index (CCI) [15]. HRQoL was assessed using the validated Spanish versions of the Saint George’s Respiratory Questionnaire (SGRQ) [16] and the COPD assessment Test (CAT) [17]. Limitation of daily living was measured with The London Chest Activity of Daily Living scale (LCADL) [18].

Pulmonary function tests included forced spirometry and body plethysmography, and measurements of carbon monoxide diffusing capacity (DLCO). These tests were performed in accordance with the standards of the European Respiratory Society [19,20]. For theoretical values, we considered those of the European Respiratory Society [20].

FC was evaluated by the 6MWT total distance in meters and PA was quantified by the average number of steps per day. Two 6-minute walk tests (6MWTs) were performed as per American Thoracic Society guidelines [21]. Physical activity was measured using an accelerometer (SenseWear Pro 3 Body Media Inc, Pittsburgh, PA, USA). Participants kept the armband on over 7 consecutive days at all times except when performing daily personal hygiene. Extension of the dominant quadriceps was performed using a hand-held dynamometer (Lafayette hand-held dynamometer model 01163).

### Follow-up

Participants underwent a follow-up assessment up to 3 times after joining the study, with planned annual evaluations having finally a median follow-up of 50 months (interquartile range, 31-62 months), due to the fact that some patients were not able to assist, or were not stable, to visits exactly every 12 months. No interventions were performed related to this study, and the research team did not take part in patients’ routine care or the treatment of any exacerbations.

Participants’ hospital medical records on hospitalizations were reviewed at each assessment during the follow-up period. Vital status was established by reviewing the hospital database and public death registries.

### Statistical analysis

PA was quantified by the average number of steps per day, and FC was evaluated by the 6MWT total distance in meters. The two were dichotomized separately for men and women. There are no previous studies that look at the relationship between FC and PA and HRQoL in COPD patients. Therefore, we had to hypothesize that individuals in the new CanDo group and those in the CańtDo group may have a difference of 10 units in the total scores of the St Georgés Respiratory Questionnaire (assuming a pooled standard deviation of 15 units). We also assumed a different sample size between groups being larger the CanDo group in relation to the CańtDo group (ratio 4:1). Based on those numbers, and to achieve a power of 80% and a level of significance of 5% (two sided), the study would require a sample size of 23 patients for the CańtDo group and 92 for the CanDo group (i.e. a total sample size of 115; to ensure that the reference group is 4 times larger than the test group) for detecting a true difference in means between both groups. To select the optimal threshold for dichotomizing PA and FC, logistic regression models were adjusted separately for men, women, FC and PA, with mortality over the course of the study as a response variable, in a similar fashion to that used in the study by Vaes et al. [11]. We considered cut-off points corresponding to the point closest to the top-left part of the Receiver Operating Characteristic (ROC) curve to be optimal. The *pROC* package in R was used to estimate these cut-off points [22]. Using these threshold values, men and women were classified into the following “CanDo/DoDo” quadrants (as in [11]): 1. Can/Do, in which both FC and PA were preserved; 2. Can/Don’t with FC preserved but low PA, 3. Can’t/Do with low FC but preserved PA, and 4. Can’t/Don’t with low FC and low PA. For purposes of illustration, the FC and PA values have been centered on the threshold value so that males and females are drawn together in the quadrants. Differences between the quadrants were analyzed using Kruskal Wallis and Fisher’s exact test methods.

Different outcomes were considered with regard to the four PA/FC categories described above. We first addressed the patient’s HRQoL at baseline. In particular, SGRQ [16] and CAT [17] were modeled considering beta-binomial regression models using the PROreg package of R [23]. Secondly, we studied the probability of hospital admissions and the number of hospitalizations in one year after patients joined the study, modeled by means of a logistic and Poisson regression model, respectively. Using a logistic regression model, we analyzed “1-year Mortality” since admission. Finally, we studied the probability of survival during the study period by means of a Cox proportional hazard regression model (Survival Time). Univariate models were fitted with “CanDo/DoDo” quadrants for all outcomes of interest as the explanatory variable. In addition, in order to consider the effect of potential confounder, multivariate models were fitted where the effect of the quadrants was measured and adjusted by other statistically significant clinical and sociodemographic covariates. All effects were considered significant at a significant level of alpha=0.05. All the analyses were performed using the software R version 4.3.2 [24].

## RESULTS

Four hundred and one consecutive patients were included in this study. The optimal thresholds obtained to classify PA were 4,526 and 4,509 average steps per day for men and women, respectively. The optimal thresholds obtained to classify FC were 463 and 456 meters for men and women, respectively. The main characteristics of the sample are shown in Table 1. The distribution of the patients in the FC/PA grid is showed in Figure 1.

**Figure 1.**
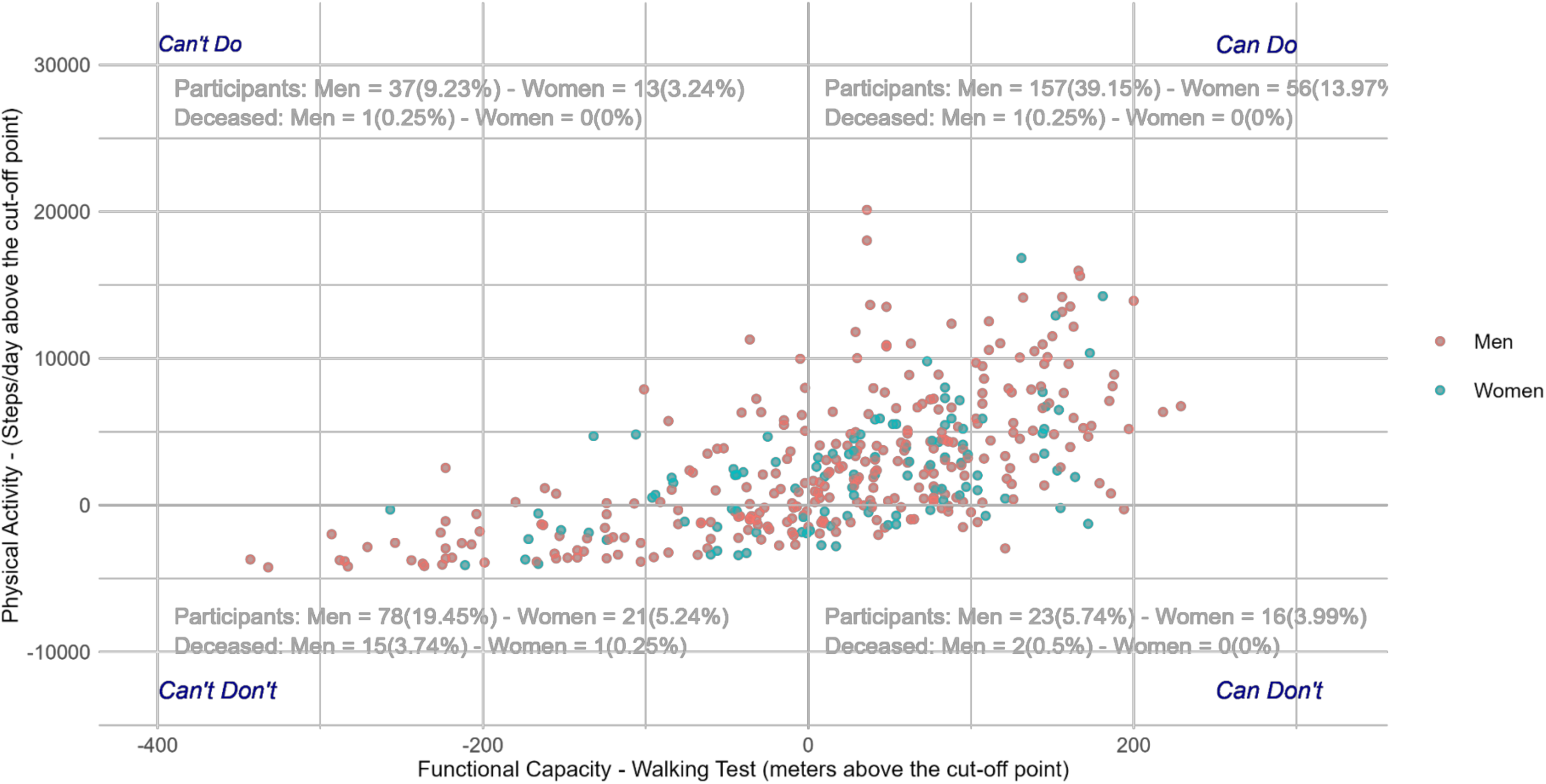
The graphical representation illustrates the distribution of patients (colored by gender) along the Can Do, Do Do quadrants. Functional Capacity and Physical Activity are both centered at the corresponding cut-off points: 463 meters and 4526 steps per day for men, and 456 meters and 4509 steps per day for women. Therefore, each point in the graphic reports the number of meters and steps per day above these cut-off points. Additionally, the distribution of participants and deceased individuals, along with their respective percentages (depicted in grey text), is delineated within each quadrant: Can’t Do and Can Do (upper section) and Can’t Don’t and Can Don’t (lower section).

**Table 1.**
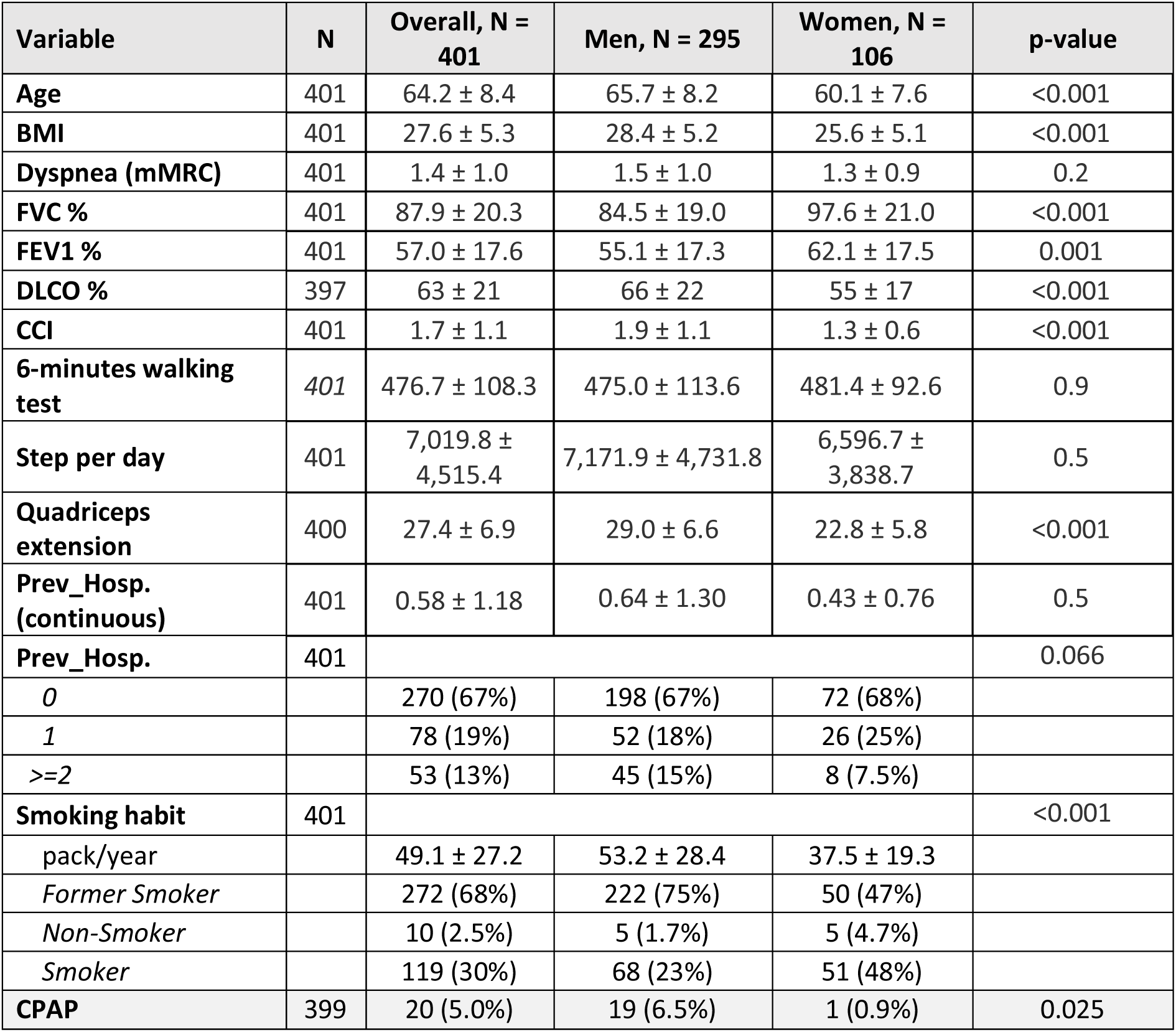
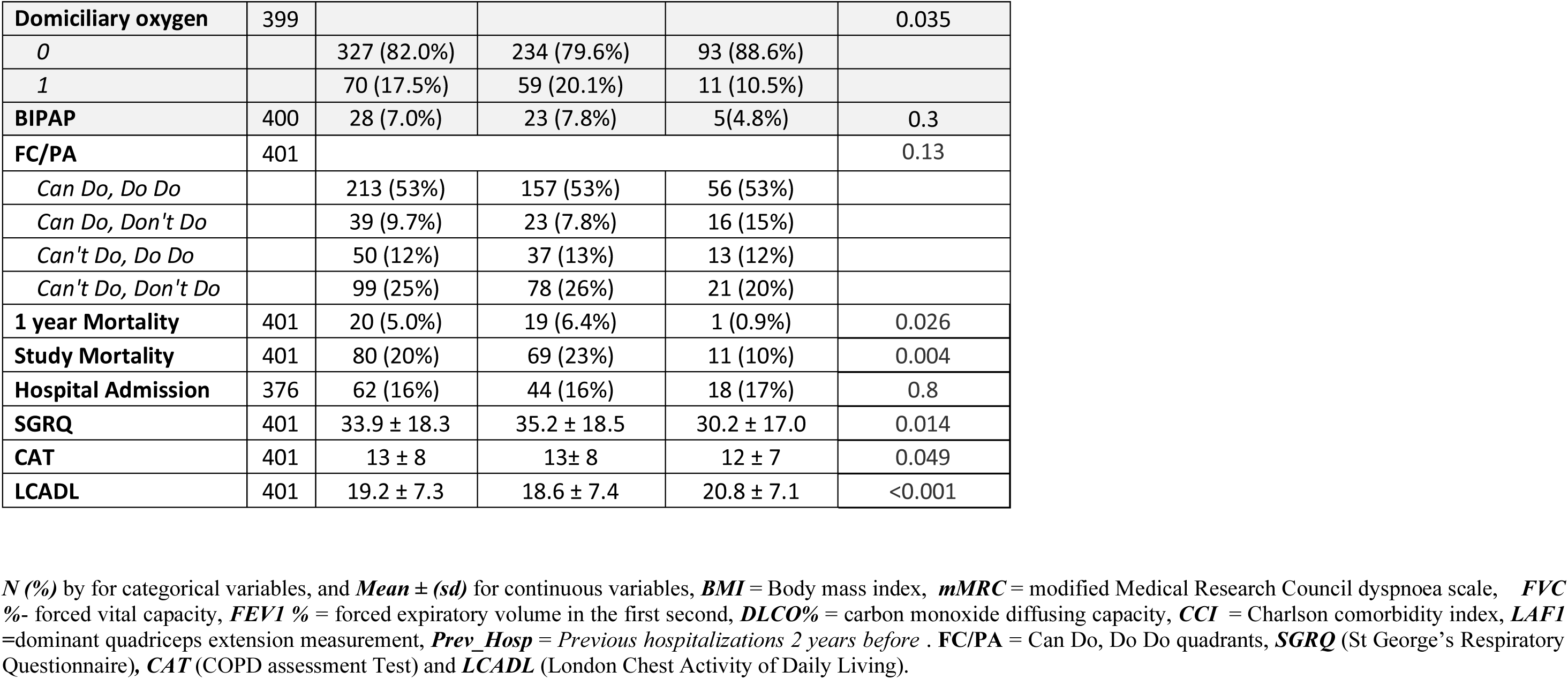
Descriptive sociodemographic and clinical characteristics of the sample.

| Variable | N | Overall, N = 401 | Men, N = 295 | Women, N = 106 | p-value |
| --- | --- | --- | --- | --- | --- |
| Age | 401 | 64.2 ± 8.4 | 65.7 ± 8.2 | 60.1 ± 7.6 | <0.001 |
| BMI | 401 | 27.6 ± 5.3 | 28.4 ± 5.2 | 25.6 ± 5.1 | <0.001 |
| Dyspnea (mMRC) | 401 | 1.4 ± 1.0 | 1.5 ± 1.0 | 1.3 ± 0.9 | 0.2 |
| FVC % | 401 | 87.9 ± 20.3 | 84.5 ± 19.0 | 97.6 ± 21.0 | <0.001 |
| FEV1 % | 401 | 57.0 ± 17.6 | 55.1 ± 17.3 | 62.1 ± 17.5 | 0.001 |
| DLCO % | 397 | 63 ± 21 | 66 ± 22 | 55 ± 17 | <0.001 |
| CCI | 401 | 1.7 ± 1.1 | 1.9 ± 1.1 | 1.3 ± 0.6 | <0.001 |
| 6-minutes walking test | 401 | 476.7 ± 108.3 | 475.0 ± 113.6 | 481.4 ± 92.6 | 0.9 |
| Step per day | 401 | 7,019.8 ± 4,515.4 | 7,171.9 ± 4,731.8 | 6,596.7 ± 3,838.7 | 0.5 |
| Quadriceps extension | 400 | 27.4 ± 6.9 | 29.0 ± 6.6 | 22.8 ± 5.8 | <0.001 |
| Prev_Hosp. (continuous) | 401 | 0.58 ± 1.18 | 0.64 ± 1.30 | 0.43 ± 0.76 | 0.5 |
| Prev_Hosp. | 401 |  |  |  | 0.066 |
| 0 |  | 270 (67%) | 198 (67%) | 72 (68%) |  |
| 1 |  | 78 (19%) | 52 (18%) | 26 (25%) |  |
| >=2 |  | 53 (13%) | 45 (15%) | 8 (7.5%) |  |
| Smoking habit | 401 |  |  |  | <0.001 |
| pack/year |  | 49.1 ± 27.2 | 53.2 ± 28.4 | 37.5 ± 19.3 |  |
| Former Smoker |  | 272 (68%) | 222 (75%) | 50 (47%) |  |
| Non-Smoker |  | 10 (2.5%) | 5 (1.7%) | 5 (4.7%) |  |
| Smoker |  | 119 (30%) | 68 (23%) | 51 (48%) |  |
| CPAP | 399 | 20 (5.0%) | 19 (6.5%) | 1 (0.9%) | 0.025 |
| **Domiciliary oxygen** | 399 |  |  |  | 0.035 |
| 0 |  | 327 (82.0%) | 234 (79.6%) | 93 (88.6%) |  |
| 1 |  | 70 (17.5%) | 59 (20.1%) | 11 (10.5%) |  |
| **BIPAP** | 400 | 28 (7.0%) | 23 (7.8%) | 5(4.8%) | 0.3 |
| **FC/PA** | 401 |  |  |  | 0.13 |
| Can Do, Do Do |  | 213 (53%) | 157 (53%) | 56 (53%) |  |
| Can Do, Don't Do |  | 39 (9.7%) | 23 (7.8%) | 16 (15%) |  |
| Can't Do, Do Do |  | 50 (12%) | 37 (13%) | 13 (12%) |  |
| Can't Do, Don't Do |  | 99 (25%) | 78 (26%) | 21 (20%) |  |
| **1 year Mortality** | 401 | 20 (5.0%) | 19 (6.4%) | 1 (0.9%) | 0.026 |
| **Study Mortality** | 401 | 80 (20%) | 69 (23%) | 11 (10%) | 0.004 |
| **Hospital Admission** | 376 | 62 (16%) | 44 (16%) | 18 (17%) | 0.8 |
| **SGRQ** | 401 | 33.9 ± 18.3 | 35.2 ± 18.5 | 30.2 ± 17.0 | 0.014 |
| **CAT** | 401 | 13 ± 8 | 13± 8 | 12 ± 7 | 0.049 |
| **LCADL** | 401 | 19.2 ± 7.3 | 18.6 ± 7.4 | 20.8 ± 7.1 | <0.001 |

*N (%)* by for categorical variables, and *Mean ± (sd)* for continuous variables, *BMI* = Body mass index, *mMRC* = modified Medical Research Council dyspnoea scale, *FVC* %- forced vital capacity, *FEV1* % = forced expiratory volume in the first second, *DLCO*% = carbon monoxide diffusing capacity, *CCI* = Charlson comorbidity index, *LAFI* =dominant quadriceps extension measurement, *Prev\_Hosp* = *Previous hospitalizations 2 years before* . *FC/PA* = Can Do, Do Do quadrants, *SGRQ* (St George's Respiratory Questionnaire), *CAT* (COPD assessment Test) and *LCADL* (London Chest Activity of Daily Living).

Table 2 shows the four FC/PA categories, the “Can/Do” group had the best results and outcomes. On the contrary, the worst clinical situation and outcomes were for the “Can’t/Don’t” group.

**Table 2.**
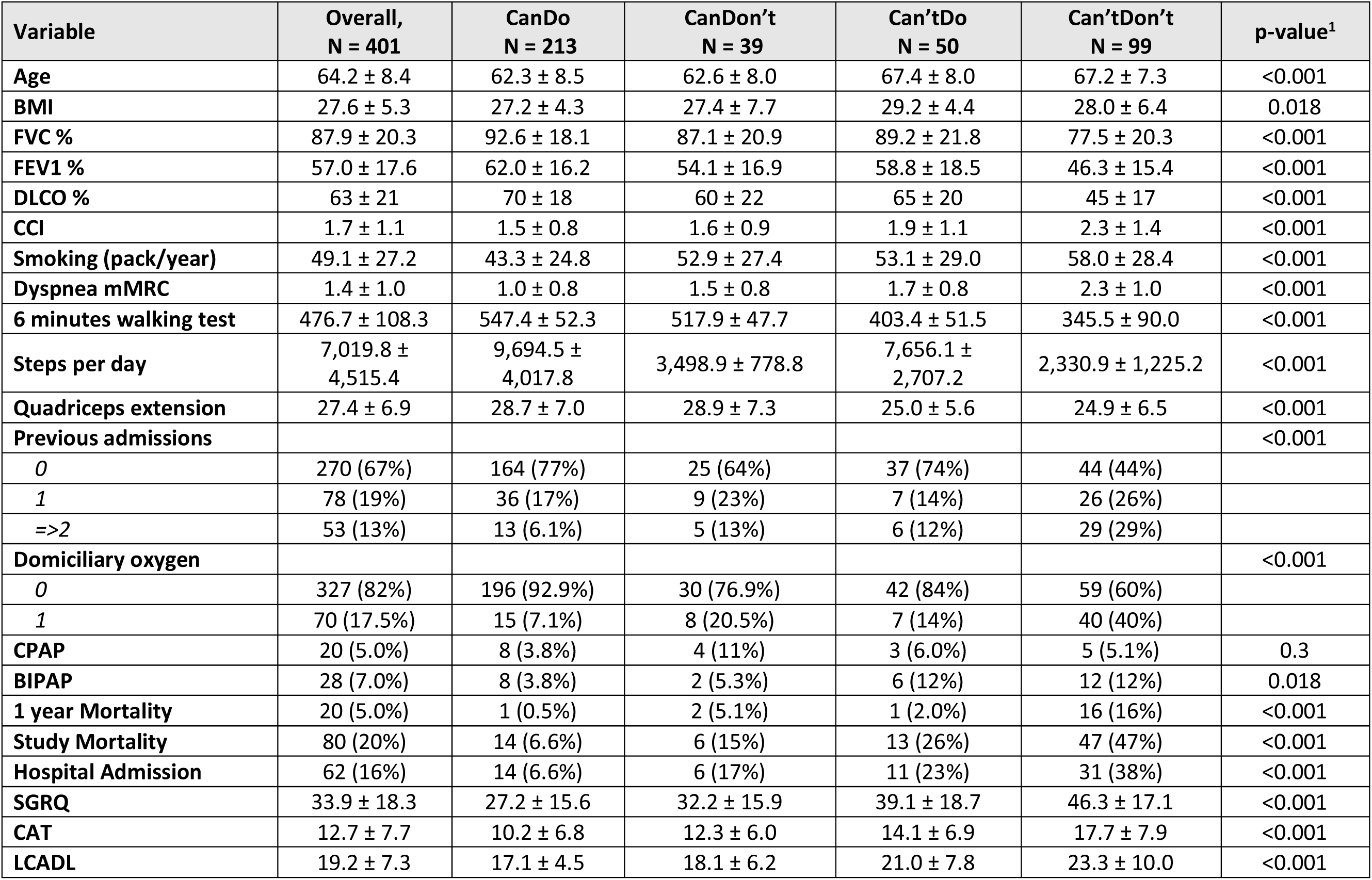
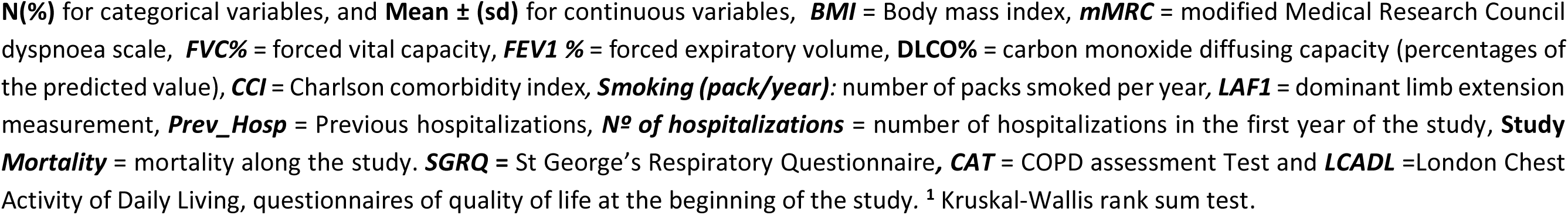
Patient characteristics per “CanDo-DoDo” quadrants.

| Variable | Overall,<br>N = 401 | CanDo<br>N = 213 | CanDon't<br>N = 39 | Can'tDo<br>N = 50 | Can'tDon't<br>N = 99 | p-value<sup>1</sup> |
| --- | --- | --- | --- | --- | --- | --- |
| Age | 64.2 ± 8.4 | 62.3 ± 8.5 | 62.6 ± 8.0 | 67.4 ± 8.0 | 67.2 ± 7.3 | <0.001 |
| BMI | 27.6 ± 5.3 | 27.2 ± 4.3 | 27.4 ± 7.7 | 29.2 ± 4.4 | 28.0 ± 6.4 | 0.018 |
| FVC % | 87.9 ± 20.3 | 92.6 ± 18.1 | 87.1 ± 20.9 | 89.2 ± 21.8 | 77.5 ± 20.3 | <0.001 |
| FEV1 % | 57.0 ± 17.6 | 62.0 ± 16.2 | 54.1 ± 16.9 | 58.8 ± 18.5 | 46.3 ± 15.4 | <0.001 |
| DLCO % | 63 ± 21 | 70 ± 18 | 60 ± 22 | 65 ± 20 | 45 ± 17 | <0.001 |
| CCI | 1.7 ± 1.1 | 1.5 ± 0.8 | 1.6 ± 0.9 | 1.9 ± 1.1 | 2.3 ± 1.4 | <0.001 |
| Smoking (pack/year) | 49.1 ± 27.2 | 43.3 ± 24.8 | 52.9 ± 27.4 | 53.1 ± 29.0 | 58.0 ± 28.4 | <0.001 |
| Dyspnea mMRC | 1.4 ± 1.0 | 1.0 ± 0.8 | 1.5 ± 0.8 | 1.7 ± 0.8 | 2.3 ± 1.0 | <0.001 |
| 6 minutes walking test | 476.7 ± 108.3 | 547.4 ± 52.3 | 517.9 ± 47.7 | 403.4 ± 51.5 | 345.5 ± 90.0 | <0.001 |
| Steps per day | 7,019.8 ±<br>4,515.4 | 9,694.5 ±<br>4,017.8 | 3,498.9 ± 778.8 | 7,656.1 ±<br>2,707.2 | 2,330.9 ± 1,225.2 | <0.001 |
| Quadriceps extension | 27.4 ± 6.9 | 28.7 ± 7.0 | 28.9 ± 7.3 | 25.0 ± 5.6 | 24.9 ± 6.5 | <0.001 |
| Previous admissions |  |  |  |  |  | <0.001 |
| 0 | 270 (67%) | 164 (77%) | 25 (64%) | 37 (74%) | 44 (44%) |  |
| 1 | 78 (19%) | 36 (17%) | 9 (23%) | 7 (14%) | 26 (26%) |  |
| =>2 | 53 (13%) | 13 (6.1%) | 5 (13%) | 6 (12%) | 29 (29%) |  |
| Domiciliary oxygen |  |  |  |  |  | <0.001 |
| 0 | 327 (82%) | 196 (92.9%) | 30 (76.9%) | 42 (84%) | 59 (60%) |  |
| 1 | 70 (17.5%) | 15 (7.1%) | 8 (20.5%) | 7 (14%) | 40 (40%) |  |
| CPAP | 20 (5.0%) | 8 (3.8%) | 4 (11%) | 3 (6.0%) | 5 (5.1%) | 0.3 |
| BIPAP | 28 (7.0%) | 8 (3.8%) | 2 (5.3%) | 6 (12%) | 12 (12%) | 0.018 |
| 1 year Mortality | 20 (5.0%) | 1 (0.5%) | 2 (5.1%) | 1 (2.0%) | 16 (16%) | <0.001 |
| Study Mortality | 80 (20%) | 14 (6.6%) | 6 (15%) | 13 (26%) | 47 (47%) | <0.001 |
| Hospital Admission | 62 (16%) | 14 (6.6%) | 6 (17%) | 11 (23%) | 31 (38%) | <0.001 |
| SGRQ | 33.9 ± 18.3 | 27.2 ± 15.6 | 32.2 ± 15.9 | 39.1 ± 18.7 | 46.3 ± 17.1 | <0.001 |
| CAT | 12.7 ± 7.7 | 10.2 ± 6.8 | 12.3 ± 6.0 | 14.1 ± 6.9 | 17.7 ± 7.9 | <0.001 |
| LCADL | 19.2 ± 7.3 | 17.1 ± 4.5 | 18.1 ± 6.2 | 21.0 ± 7.8 | 23.3 ± 10.0 | <0.001 |

**N(%)** for categorical variables, and **Mean ± (sd)** for continuous variables, **BMI** = Body mass index, **mMRC** = modified Medical Research Council dyspnoea scale, **FVC%** = forced vital capacity, **FEV1 %** = forced expiratory volume, **DLCO%** = carbon monoxide diffusing capacity (percentages of the predicted value), **CCI** = Charlson comorbidity index, **Smoking (pack/year)**: number of packs smoked per year, **LAF1** = dominant limb extension measurement, **Prev\_Hosp** = Previous hospitalizations, **Nº of hospitalizations** = number of hospitalizations in the first year of the study, **Study Mortality** = mortality along the study. **SGRQ** = St George's Respiratory Questionnaire, **CAT** = COPD assessment Test and **LCADL** = London Chest Activity of Daily Living, questionnaires of quality of life at the beginning of the study. <sup>1</sup> Kruskal-Wallis rank sum test.

Table 3 shows the univariate analysis for the “CanDo/DoDo” quadrants for clinical outcomes, and Table 4 shows the estimates obtained for the univariate models for HRQoL outcomes. There were statistically significant differences between the “Can/Do” and “Can’t/Don’t” groups for all clinical and HRQoL outcomes. Nevertheless, no significant differences were observed for basal LCADL. Figure 2 gives the Kaplan-Meier survival estimation curves for the four “CanDo/DoDo” quadrants, showing that the “Can’t/Don’t” group had the poorest survival rate, followed by the “Can’t/Do” group until three years follow-up.

**Figure 2.**
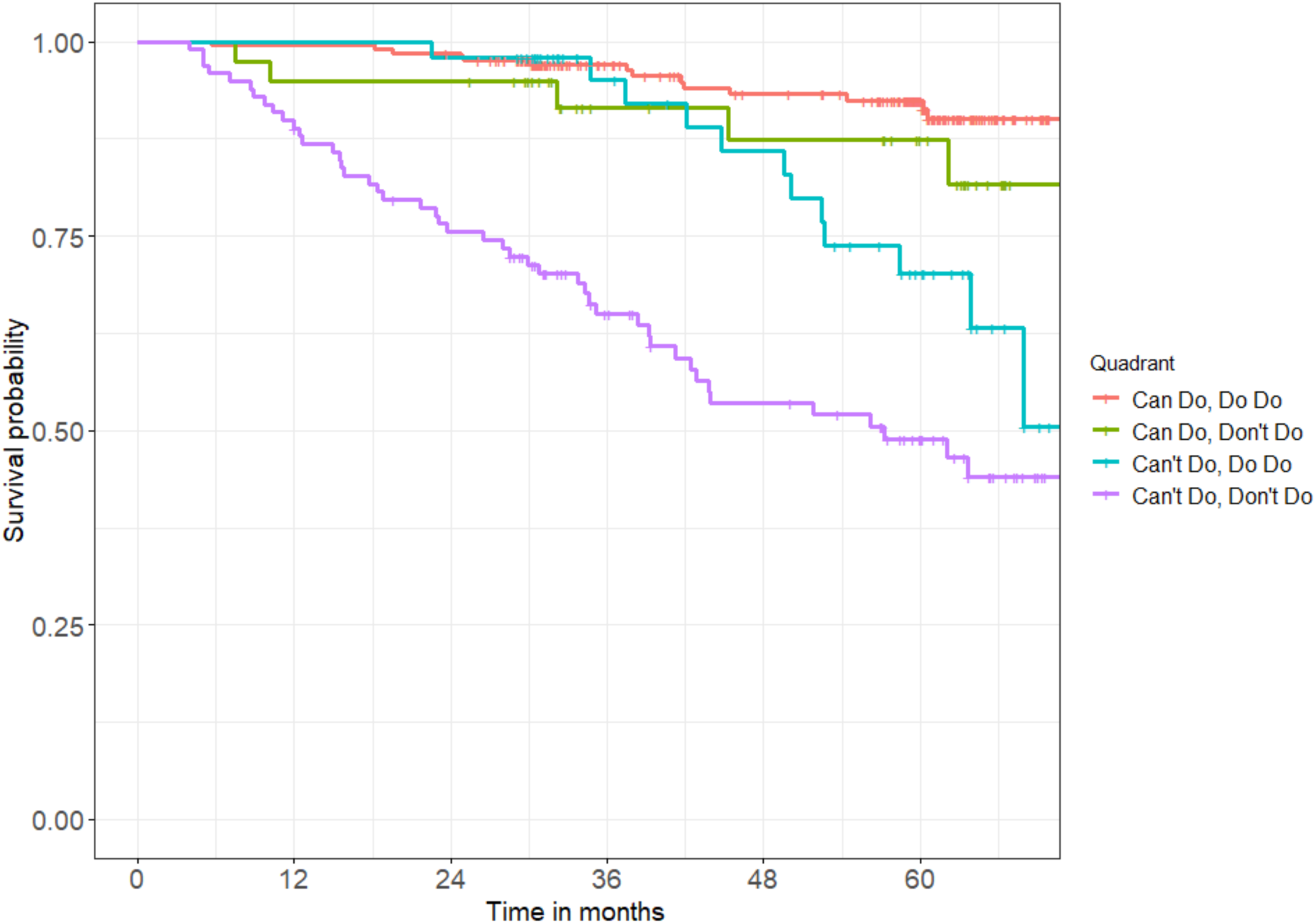
Kaplan-Meier survival estimation curves for each of the Can Do, Do Do quadrants.

**Table 3.** Univariate models considering “CanDo-DoDo” quadrants as covariate for outcomes I.

| Variable | 1 year Mortality |  | Survival |  | Hospital Admission |  | Number of hospitalizations |  |
| --- | --- | --- | --- | --- | --- | --- | --- | --- |
|  | OR<br>(CI-95%) | p-value | HR<br>(CI-95%) | p-value | OR<br>(CI-95%) | p-value | IRR<br>(CI-95%) | p-value |
| **FC/PA** |  |  |  |  |  |  |  |  |
| *Can Do, Do Do* | — |  | — |  | — |  | — |  |
| *Can't Do, Do Do* | 4.33<br>(0.17, 111) | 0.3 | 4.24<br>(1.99, 9.03) | <0.001 | 4.3<br>(1.78, 10.2) | <0.001 | 3.71<br>(1.70, 8.17) | <0.001 |
| *Can Do, Don't Do* | 11.5<br>(1.07, 250) | 0.05 | 2.57<br>(0.99, 6.70) | 0.053 | 2.81<br>(0.94, 7.62) | 0.049 | 2.04<br>(0.74, 5.32) | 0.15 |
| *Can't Do, Don't Do* | 40.9<br>(8.14, 743) | <0.001 | 9.89<br>(5.43, 18.0) | <0.001 | 8.55<br>(4.32, 17.7) | <0.001 | 6.04<br>(3.27, 11.5) | <0.001 |
| **AUC \***<br>**(CI-95%)** | 0.83 (0.76,0.90) |  | 0.76 (0.71,0.81) |  | 0.73 (0.67,0.80) |  | 0.4 |  |
| **AIC** | 133.88 |  |  |  | 303.34 |  | 457.8 |  |
| **Deviance** | 125.88 |  |  |  | 295.34 |  | 189.5 |  |

Outcomes: 1 year mortality, risk of hospitalization and number of hospitalizations in one year period, and 3-years survival. **OR** = odds ratios; **HR** = hazard ratios. **IRR** = Incidence Rate Ratio. \* For the Cox proportional hazards model (survival) the c-index is used, and for the Poisson model (number of hospitalizations) the Theta parameter is used.

**Table 4.** Univariate models considering “CanDo-DoDo” quadrants as covariates for the outcomes II.

| Variable | SGRQ |  | CAT |  | LCADL |  |
| --- | --- | --- | --- | --- | --- | --- |
|  | OR (CI-95%) | p-value | OR (CI-95%) | p-value | OR (CI-95%) | p-value |
| **FC/PA** |  |  |  |  |  |  |
| *Can Do, Do Do* | — | — | — | — | — | — |
| *Can't Do, Do Do* | 1.74 (1.37-2.21) | <0.001 | 2.06 (1.37-3.09) | <0.001 | 1.34 (1.15,1.56) | <0.001 |
| *Can Do, Don't Do* | 1.32 (1.01-1.73) | 0.04 | 1.57 (0.99-2.53) | 0.1 | 1.09 (0.91,1.30) | 0.37 |
| *Can't Do, Don't Do* | 2.39 (1.99-2.87) | <0.001 | 3.31 (2.43-4.51) | <0.001 | 1.58 (1.41,1.78) | <0.001 |
| *log(phi)* | -2.22 | — | -839 | — | -5.242 | — |
| *Std. Error* | 0.098 | — | — | — | 0.6 | — |
| *Deviance* | 2230.6 | — | 821.53 | — | 1774.07 | — |

Outcomes: quality of life at the beginning of the study . **SGRQ** = St George’s Respiratory Questionnaire, **CAT** = COPD assessment Test and **LCADL** =London Chest Activity of Daily Living. **OR** = odds ratios; **HR** = hazard ratios. **IRR** = Incidence Rate Ratio.

In the multivariate model of mortality in the first year, taking the “Can/Do” group as a reference, the “Can’t/Do” and “Can’t/Don’t” groups had a higher risk of dying in one year, also adjusting by the CCI (AUC: 0.88). The Cox proportional hazard model, also showed that the “Can’t/Do” and the “Can’t/Don’t” had a higher HR of dying, including as additional predictors the CCI (HR 1.18), age (HR 1.10), and FEV1% (HR 0.97) and DLCO (HR 0.98) with lower risk for higher values (c-index: 0.83). In the model of risk of hospital admission, the “Can’t/Do” and the “Can’t/Don’t” groups had the highest risk of admission, in addition to the following variables: hospitalization in the two years prior to joining the study (2 or more OR 3.14), being a woman (OR 2.86) and being older (OR 1.07), while having less risk with higher FEV1% (OR 0.95) (AUC: 0.84). (Table 5) Worse SGRQ scores at the beginning of the study were related to the “Can’t/Do” and the “Can’t/Don’t” groups while being a woman (OR 0.83), older age (OR 0.99) and higher FEV1% (OR 0.99) had better scores. In the model of CAT prediction, the “Can’t/Do” and the “Can’t/Don’t” groups again had worse scores as well as hospitalizations in the two years before inclusion in the study (2 or more OR 1.94), while being a woman (OR 0.70) and being older (OR 0.96) gave better CAT scores. In the model of LCADL scores too, the “Can’t/Do” and the “Can’t/Don’t” groups had worse scores as well as older age (OR 1.27) and have had more prior hospitalizations (2 or more OR 1.26), while having higher FEV1% (OR 0.99) resulted in better scores. (Table 6)

**Table 5.**
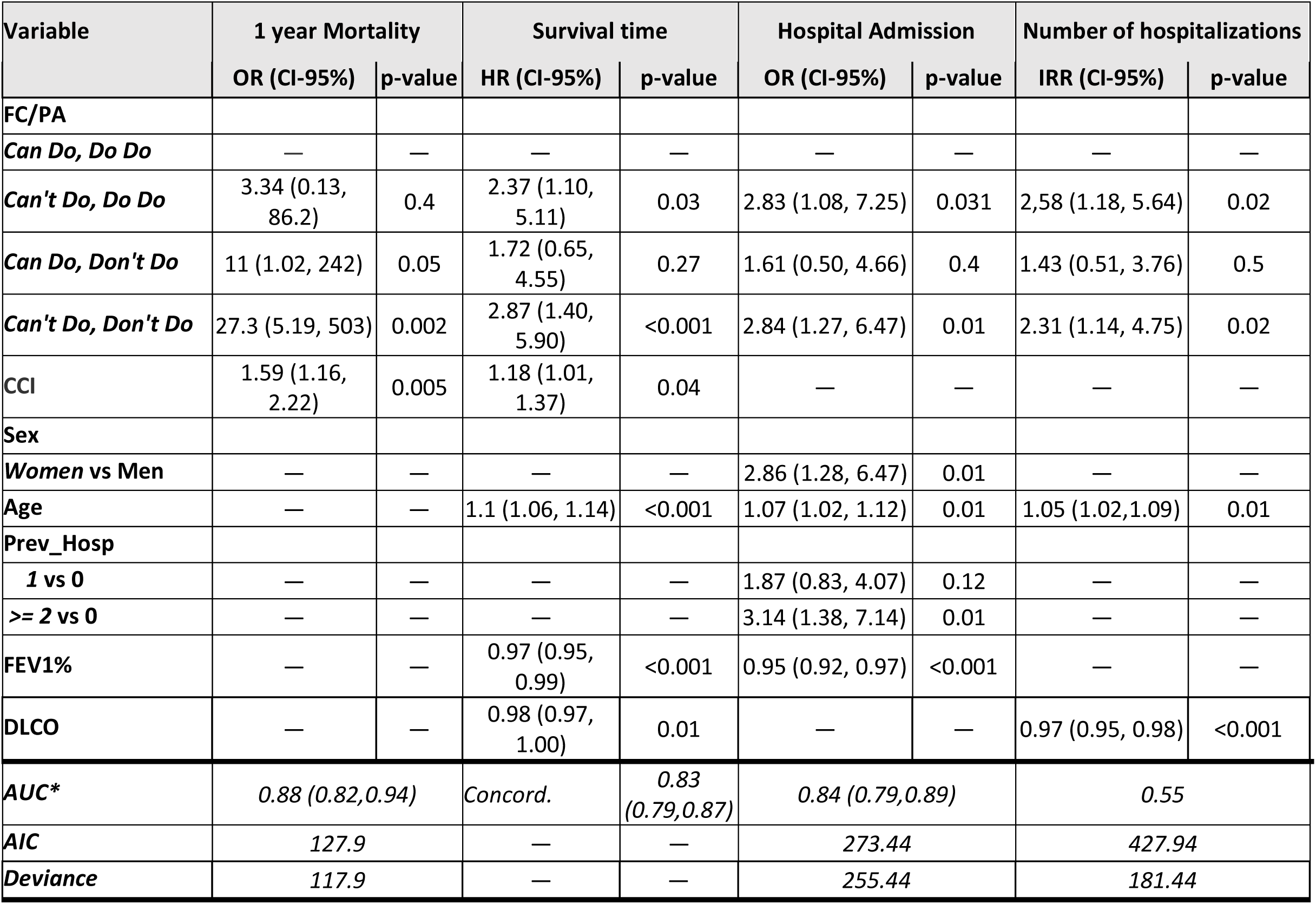
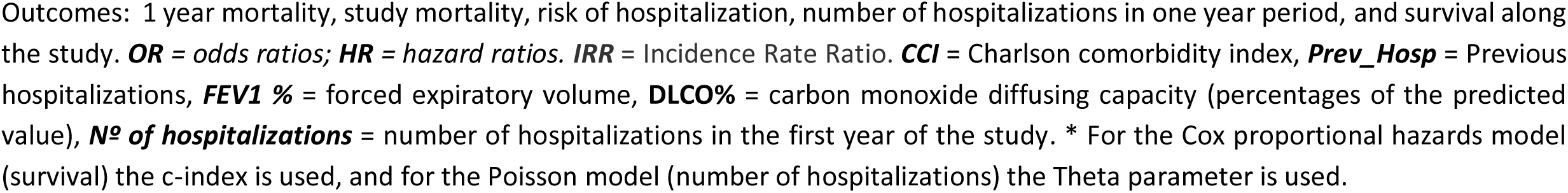
Multivariate models I.

| Variable | 1 year Mortality |  | Survival time |  | Hospital Admission |  | Number of hospitalizations |  |
| --- | --- | --- | --- | --- | --- | --- | --- | --- |
|  | OR (CI-95%) | p-value | HR (CI-95%) | p-value | OR (CI-95%) | p-value | IRR (CI-95%) | p-value |
| **FC/PA** |  |  |  |  |  |  |  |  |
| *Can Do, Do Do* | — | — | — | — | — | — | — | — |
| *Can't Do, Do Do* | 3.34 (0.13, 86.2) | 0.4 | 2.37 (1.10, 5.11) | 0.03 | 2.83 (1.08, 7.25) | 0.031 | 2,58 (1.18, 5.64) | 0.02 |
| *Can Do, Don't Do* | 11 (1.02, 242) | 0.05 | 1.72 (0.65, 4.55) | 0.27 | 1.61 (0.50, 4.66) | 0.4 | 1.43 (0.51, 3.76) | 0.5 |
| *Can't Do, Don't Do* | 27.3 (5.19, 503) | 0.002 | 2.87 (1.40, 5.90) | <0.001 | 2.84 (1.27, 6.47) | 0.01 | 2.31 (1.14, 4.75) | 0.02 |
| **CCI** | 1.59 (1.16, 2.22) | 0.005 | 1.18 (1.01, 1.37) | 0.04 | — | — | — | — |
| **Sex** |  |  |  |  |  |  |  |  |
| *Women vs Men* | — | — | — | — | 2.86 (1.28, 6.47) | 0.01 | — | — |
| **Age** | — | — | 1.1 (1.06, 1.14) | <0.001 | 1.07 (1.02, 1.12) | 0.01 | 1.05 (1.02,1.09) | 0.01 |
| **Prev_Hosp** |  |  |  |  |  |  |  |  |
| *1 vs 0* | — | — | — | — | 1.87 (0.83, 4.07) | 0.12 | — | — |
| *>= 2 vs 0* | — | — | — | — | 3.14 (1.38, 7.14) | 0.01 | — | — |
| **FEV1%** | — | — | 0.97 (0.95, 0.99) | <0.001 | 0.95 (0.92, 0.97) | <0.001 | — | — |
| **DLCO** | — | — | 0.98 (0.97, 1.00) | 0.01 | — | — | 0.97 (0.95, 0.98) | <0.001 |
| **AUC\*** | 0.88 (0.82,0.94) |  | Concord. | 0.83 (0.79,0.87) | 0.84 (0.79,0.89) |  | 0.55 |  |
| **AIC** | 127.9 |  | — | — | 273.44 |  | 427.94 |  |
| **Deviance** | 117.9 |  | — | — | 255.44 |  | 181.44 |  |

Outcomes: 1 year mortality, study mortality, risk of hospitalization, number of hospitalizations in one year period, and survival along the study. **OR** = odds ratios; **HR** = hazard ratios. **IRR** = Incidence Rate Ratio. **CCI** = Charlson comorbidity index, **Prev\_Hosp** = Previous hospitalizations, **FEV1 %** = forced expiratory volume, **DLCO%** = carbon monoxide diffusing capacity (percentages of the predicted value), **Nº of hospitalizations** = number of hospitalizations in the first year of the study. \* For the Cox proportional hazards model (survival) the c-index is used, and for the Poisson model (number of hospitalizations) the Theta parameter is used.

**Table 6.** Multivariate models II.

| Variable | SGRQ |  | CAT |  | LCADL |  |
| --- | --- | --- | --- | --- | --- | --- |
|  | OR (CI-95%) | p-value | OR (CI-95%) | p-value | OR (CI-95%) | p-value |
| **FC/PA** |  |  |  |  |  |  |
| *Can Do, Do Do* | — |  | — |  |  |  |
| *Can't Do, Do Do* | 1.83 (1.45-2.32) | <0.001 | 2.44 (1.60-3.71) | <0.001 | 1.31 (1.13-1.52) | 0.001 |
| *Can Do, Don't Do* | 1.25 (0.96-1.62) | 0.1 | 1.62 (1.00-2.61) | 0.05 | 0.996 (1.25-1.61) | 0.99 |
| *Can't Do, Don't Do* | 2.15 (1.76-2.61) | <0.001 | 3.39 (2.40-4.79) | <0.001 | 1.42 (0.84-1.19) | <0.001 |
| **Sex** |  |  |  |  |  |  |
| *Men* | — |  | — |  |  |  |
| *Women* | 0.83 (0.69-1.00) | 0.04 | 0.7 (0.50-0.98) | 0.04 |  |  |
| **Age** | 0.99 (0.98-0.99) | <0.001 | 0.96 (0.95-0.98) | <0.001 | 1.272 (1.14-1.42) | <0.001 |
| **Prev_Hosp** |  |  |  |  |  |  |
| *0* | — |  | — |  |  |  |
| *1* | — |  | 1.12 (0.78-1.59) | 0.55 | 0.97 (0.85-1.10) | 0.62 |
| *>= 2* | — |  | 1.94 (1.31-2.86) | <0.001 | 1.26 (1.09-1.46) | 0.002 |
| **FEV1%** | 0.99 (0.98-0.99) | <0.001 | — |  | 0.995 (0.99-1.00) | 0.001 |
| **log(phi)** | -2.39 |  | -839 |  | -6.53 |  |
| **Deviance** | 2184.99 |  | 795.13 |  | 1709.78 |  |

Outcomes: **SGRQ** = St George's Respiratory Questionnaire, **CAT** = COPD assessment Test and **LCADL** =London Chest Activity of Daily Living questionnaires. **OR** = odds ratios; **CCI** =Charlson comorbidity index, **Prev\_Hosp** = Previous hospitalizations, **FEV1 %**- forced expiratory volume.

## DISCUSSION

This observational study highlights: 1. The importance of FC and PA as prognosis factors; 2. Their relationship with HRQoL and daily life limitations; 3. Their potential usefulness as tools for classifying people with COPD, as proposed by Vaes et al. [11], in order to design interventions to improve these outcomes.

In addition to the classical emphasis on hospitalizations, HRQoL and mortality as key outcomes in people with COPD, it is also critically important to focus on maintaining maximal FC and thus the essential goal of an independent life over time. It is also necessary to keep in mind that a decline in FC is part of the ageing process and this effect is accelerated in chronic diseases. All of these elements form an essential domain of the “intrinsic capacity” construct [25].

It is useful to look at the trajectories of PA and FC in the general population. The walking distance of American adults halves between the ages of 40 and 70, associated with a decrease in walking speed (33% slower in the 60s than in the 40s). From an evolutionary perspective, our hunter-gatherer predecessors displayed a much smaller decline in these parameters over time starting from a much higher daily level of PA [26].

Studies of top athletes have shown that the time they require to perform certain disciplines follows a linear decline with age, with a break-point at the seventh/eighth decade after which there is an exponential increase in the times taken. This may reflect a decline in physiological FC directly related to ageing [27].

Of course, these figures relate to healthy, well-trained individuals and to our hunter-gatherer ancestors. However, they are useful for understanding the contribution of FC-loss related exclusively to ageing as it translates to patients with chronic diseases. Thus, among people with COPD an early breakdown of multi-system physiological integrity will affect the FC trajectory. This is coupled with the influence of ageing and a modern life style prone to sedentarism, all of which further influence this decline.

To the best of our knowledge, there is no information about the long-term trajectories of PA and FC in people with COPD. What we do know about PA in COPD is that it is diminished when compared to controls (people with only chronic bronchitis) [28]. With regard to the evolution of PA, in the Copenhagen City Heart Study was found a significant tendency of all categories of PA to be reduced [29]. Moreover, Waschki et al. in a 3-year follow-up study described a decrease in PA in all stages of COPD, and especially in the more severe stages of obstruction [30].

FC, however, appears to show a different behavior. In a cohort of people with COPD followed for an average of almost 3 years, 6MWT remained stable whereas PA declined by 451 steps per year [31]. Notably, however, sustained low levels of PA were related to a progressive decline in the results of the 6MWT as compared with those who retained some level of PA [30]. These findings go some way to supporting some of the results of our study.

In our study, the two groups at either extreme of the cohort (Can/Do vs Can’t/Don’t) are clearly defined. The first group had the best respiratory (FEV1 and DLCO) and general clinical condition based on CCI and were the youngest. The second extreme group, by contrast, had the worst respiratory (FEV1 62.0% vs 46.3% and DLCO 70% vs 45%) and general clinical situation (CCI 1.5 vs 2.3) and consequently more symptoms (dyspnea mMRC 1.0 vs 2.3). Therefore, all outcomes were much worse among those who not only had the lowest FC but who performed less PA. In the Can/Do group it is probably necessary at least to maintain the level of PA in order to preserve the HRQoL level or even to try to increase FC in anticipation of the aging-related deterioration in this area [27]. Amongst the Can’t/Don’t group, an integral approach needs to be taken, with an attempt properly to evaluate all the comorbidities implicated in the clinical situation (even investigating other potential subclinical comorbidities that might have contributed to their clinical deterioration), adjust the treatment (especially the respiratory treatment) accordingly and then offer them an intensive and closely supervised rehabilitation program.

The Can’t/Do group is particularly interesting; although they are as old as Can’t/Don’t (both 67 years), they have a better clinical condition. Can’t/Do patients had better outcomes in survival than the Can’t/Don’t group. In terms of one-year mortality, this group had better results than the Can/Don’t group, although the small number of deaths among both groups may have influenced the statistical results. Can’t/Do patients carried out PA but probably need a structured program of PA to improve their FC (resistance/strength training and other specific subprograms included in a respiratory rehabilitation program).

The Can/Don’t group was as young as the Can/Do group (62 years old), and younger than Can’t/Do. Interestingly, they showed no statistical differences with the latter in FEV1, DLCO, CCI and dyspnea. Can/Don’t have enough FC to do PA so in that group the main target would be to analyze the reasons why they do not practice PA and try to modify this behavior in order to maintain their FC and try to change the trajectory of the likely deterioration in their FC and HRQoL over time [30].

In general terms, our study confirms the findings regarding mortality reported by Vaes et al. However, the multivariate model utilized in our study provided additional insights of the instrument into other significant outcomes, such as the risk of hospital admission, the frequency of admissions during the 3 years follow-up period, and their association with HRQoL and limitations in daily living. Essentially, the models revealed distinctions between two main groups: those who "Can" and those who "Can’t." This novel information enriches and reinforces the potential utility of classifying people with COPD based on their PA and FC.

In COPD, the most significant clinical outcomes currently include mortality, exacerbations leading to hospitalization, and HRQoL. Previous research has demonstrated associations between these outcomes and PA and FC (30, 32-38). Our study not only corroborates these established associations but also provides novel insights into how the combination of PA and FC (the quadrants) relates to these outcomes and impacts COPD management.

Also, as regards the multivariate analysis, several other factors appear independently associated to the outcomes. Some of them are well-known from previous studies, such as age and FEV1%, previous hospitalizations and CCI. However, others such as DLCO have seldom been included in the multivariate models. In the COPDGene study, DLCO was associated with an increase in the rate of exacerbations and worse HRQoL [39]. In our study, DLCO was associated with lower survival, greater number of hospitalizations and mortality. In the case of survival, the result was independent of FEV1. The impairment of DLCO probably reflected the presence of emphysema [40], but also impairment of pulmonary perfusion [41].

The study has some limitations. First, the cut-off points used to establish the four categories were calculated based on our sample, taking the same approach as in Vaes et al. [11] in order to facilitate the comparison between the studies; future studies should validate those cut-off points. Second, the sample size of some the four categories are small which reduces the precision of the estimations. Third, the proposed classification based on PA and FC and the ensuing proposal for intervention have not been validated (either internally or externally). Fouth, the follow-up period was only 3 years, providing a limited perspective on long-term outcomes. Vaes et al. [11] studied mortality over a follow-up period of 6 years. In our study, which includes additional outcomes, increasing the follow-up time could offer a better perspective on the general evolution of the patients. However, this study provides a snapshot of participants at a single point in time; participants’ conditions may evolve due to interventions (e.g., rehabilitation programs, changes in medications) and events (e.g., hospitalizations, new or worsening comorbidities), which can modify their FC and therefore their position on the quadrants over time. Ideally, a dynamic perspective should be brought into each evaluation to maintain or adjust the patient’s position on the grid and modify the care plan as needed.

In conclusion, based on the result of Vaes et al [11] and our study, it appears necessary to evaluate FC and PA in order to classify and expand the therapeutic options for all people with COPD. This approach aims not only to improve their current and short-term clinical situation, but also to change the future trajectory of decline in FC and PA and to move towards a consideration of healthspan in COPD.

## Data Availability

The datasets analysed during the current study are not publicly available due to the sensible nature of the information and the associated privacy issues but are available from the corresponding author on reasonable request.

## Acknowledgements

This work was supported in part by a grant from the Instituto de Salud Carlos III (PI13/02352) and the Instituto de Salud Carlos III (ISCIII) through the project "RD16/0001/0001" (Red de Investigación en Servicios de Salud en Enfermedades Crónicas) and the project “RD21CIII/0003/0017” (Red de Investigación en Cronicidad, Atención Primaria y Prevención y Promoción de la Salud) and co-funded by the European Union. The study received an unrestricted grant from Laboratorios Menarini. The work of IB, JNZ and IA was supported in part by the Basque Government’s Department of Education, Linguistic Policy and Culture (Departamento de Educación, Política Lingüística y Cultura del Gobierno Vasco), IT1456-22. The work of IB, AM and IA was supported in part by the Basque Government through the Basque Modeling Working Group’s “Mathematical Modeling Applied to Health” Project.

## Declaration section

### Availability of data and material

Data available on request from the authors.

## Declarations

### Conflict of interest

No financial, consultative, institutional and other relationships that might lead to bias or a conflict of interest exist for any of the authors of this study.

### Ethical approval

All procedures performed in studies involving human participants were in accordance with the ethical standards of the institutional and/or national research committee and with the 1964 Helsinki Declaration and its later amendments or comparable ethical standards.

The study protocol was approved by the Ethics Committee of the Hospital University of Galdakao (reference 16/2014).

Informed consent: All patients were required to provide written informed consent to participate in the study.

Artificial Intelligence or AI-assisted technologies have not been used in the writing process of this article.

## Notes

### Competing Interest Statement

The authors have declared no competing interest.

### Author Declarations

The study protocol was approved by the Ethics Committee of the Hospital University of Galdakao (reference 16/2014). All candidate patients were given detailed information about the study and all those included provided written informed consent.

